# An international, interlaboratory ring trial confirms the feasibility of an open-source, extraction-less “direct” RT-qPCR method for reliable detection of SARS-CoV-2 RNA in clinical samples

**DOI:** 10.1101/2021.04.10.21254091

**Authors:** Margaret G. Mills, Emily Bruce, Meei-Li Huang, Jessica W. Crothers, Ollivier Hyrien, Christopher A. L. Oura, Lemar Blake, Arianne Brown Jordan, Susan Hester, Leah Wehmas, Bernard Mari, Pascal Barby, Caroline Lacoux, Julien Fassy, Pablo Vial, Cecilia Vial, Jose R.W. Martinez, Olusola Olalekan Oladipo, Bitrus Inuwa, Ismaila Shittu, Clement A. Meseko, Roger Chammas, Carlos Ferreira Santos, Thiago José Dionísio, Thais Francini Garbieri, Viviane Aparecida Parisi, Maria Cassia Mendes-Correa, Anderson V. dePaula, Camila M. Romano, Luiz Gustavo Bentim Góes, Paola Minoprio, Angelica C. Campos, Marielton P. Cunha, Ana Paula P. Vilela, Tonney Nyirenda, Rajhab Sawasawa Mkakosya, Adamson S. Muula, Rebekah E. Dumm, Rebecca M. Harris, Constance A. Mitchell, Syril Pettit, Jason Botten, Keith R. Jerome

**Affiliations:** Virology Division, Department of Laboratory Medicine and Pathology, University of Washington, Seattle, Washington, USA; Division of Immunobiology, Department of Medicine, Robert Larner, M.D. College of Medicine, University of Vermont, Burlington, Vermont, USA; Vaccine and Infectious Disease Division, Fred Hutchinson Cancer Research Center, Seattle, Washington, USA; School of Veterinary Medicine, University of the West Indies, Trinidad and Tobago; Ministry of Health, Trinidad and Tobago; Office of Research and Development, U.S. Environmental Protection Agency, Research Triangle Park, North Carolina, USA; Université Côte d’Azur, CNRS, Institut de Pharmacologie Moléculaire et Cellulaire, FHU-OncoAge, Valbonne, France; Programa Hantavirus, Instituto de Ciencias e Innovación en Medicina, Facultad de Medicina Clínica Alemana Universidad del Desarrollo, Santiago, Chile; Biochemistry Division, National Veterinary Research Institute, Vom, Nigeria; Infectious and Transboundary Animal Diseases, National Veterinary Research Institute, Vom, Nigeria; Centro de Investigação Translacional em Oncologia, Departamento de Radiologia e Oncologia, Instituto do Cancer do Estado de São Paulo, Faculdade de Medicina da Universidade de São Paulo, São Paulo, Brazil; Bauru School of Dentistry, Department of Biological Sciences, University of São Paulo, Bauru, São Paulo, Brazil; Department of Infectious Diseases, Institute of Tropical Medicine of São Paulo, São Paulo, Brazil; Scientific Platform Pasteur-USP, Universidade de São Paulo, São Paulo, São Paulo, Brazil; Department of Microbiology, Biomedical Sciences Institute, University of São Paulo, São Paulo, Brazil; Department of Pathology, College of Medicine, University of Malawi, Blantyre, Malawi; Department of Public Health, College of Medicine, University of Malawi, Blantyre, Malawi; Children’s Hospital of Philadelphia, Philadelphia, Pennsylvania, USA; Health and Environmental Sciences Institute, Washington, DC, USA; Department of Microbiology and Molecular Genetics, Robert Larner, M.D. College of Medicine, University of Vermont, Burlington, Vermont, USA

## Abstract

Reverse transcription–quantitative polymerase chain reaction (RT-qPCR) is used worldwide to test and trace the spread of severe acute respiratory syndrome coronavirus 2 (SARS-CoV-2). “Extraction-less” or “direct” real time–reverse transcription polymerase chain reaction (RT-PCR) is an open-access qualitative method for SARS-CoV-2 detection from nasopharyngeal or oral pharyngeal samples with the potential to generate actionable data more quickly, at a lower cost, and with fewer experimental resources than full RT-qPCR. This study engaged 10 global testing sites, including laboratories currently experiencing testing limitations due to reagent or equipment shortages, in an international interlaboratory ring trial. Participating laboratories were provided a common protocol, common reagents, aliquots of identical pooled clinical samples, and purified nucleic acids and used their existing in-house equipment. We observed 100% concordance across laboratories in the correct identification of all positive and negative samples, with highly similar cycle threshold values. The test also performed well when applied to locally collected patient nasopharyngeal samples, provided the viral transport media did not contain charcoal or guanidine, both of which appeared to potently inhibit the RT-PCR reaction. Our results suggest that open-access, direct RT-PCR assays are a feasible option for more efficient COVID-19 coronavirus disease testing as demanded by the continuing pandemic.

## 1. Introduction

The global coronavirus disease (COVID-19) pandemic response depends on effective rollout of recently approved vaccines and the use of nonpharmaceutical interventions to slow the spread of the disease. Physical distancing supported by test-and-trace informed containment strategies has been promoted worldwide [1]. The effectiveness of testing as a containment strategy requires the implementation of accessible, affordable, reliable, and rapidly executable test methods that can meet the rapid pace of severe acute respiratory syndrome coronavirus 2 (SARS-CoV-2) transmission [2-4]. At present, this goal remains largely unmet.

The majority of regional and national health laboratories around the world rely on reverse transcription–quantitative polymerase chain reaction (RT-qPCR) SARS-CoV-2 virologic testing methods such as those developed by the World Health Organization and the U.S. Centers for Disease Control and Prevention (CDC) to support their public health programs [5, 6]. The methods themselves are robust and have proven to be useful standards for detection and reporting. However, sample processing time and a lack of supplies to support extraction as required to run this type of assay have resulted in widely reported backlogs and shortages in the United States and around the world [7]. In regions that also suffer from systemic financial and logistical challenges (e.g., Africa, the Caribbean, and South America), these hurdles will continue to consistently impair reliable procurement of consumables, support for staffing, and thus testing viability [8, 9]. Although the diversity and efficiency of commercial virologic and serologic test methods expands weekly, most public health laboratories lack the resources (human and capital) or remit to pivot to novel commercial methods.

To address these challenges, the nonprofit Health and Environmental Science Institute (HESI) convened an international network of public and academic COVID-19 testing laboratories — the Propagate Network — with the goal of collectively evaluating and disseminating practical, efficient, and impactful open-access methods for SARS-CoV-2 detection. The Propagate Network and others have identified “extraction-less” or “direct” real time–reverse transcription polymerase chain reaction (RT-PCR) as an open-access qualitative method for SARS-CoV-2 detection from nasopharyngeal samples with the potential to generate actionable data more quickly, at a lower cost, and with fewer experimental resources than full RT-qPCR [10, 11]. The method allows for detection of SARS-CoV-2 viral ribonucleic acid (RNA) with the omission of the most labor-intensive step — the RNA extraction step — and its associated extraction reagents. Published intralaboratory studies indicate that the technique is internally reproducible (with some loss of sensitivity compared to standard RT-PCR) and is effective in detecting both true negatives and positives. Notably, direct RT-qPCR remains sufficiently sensitive to detect viral RNA from patients most likely to be infectious (cycle threshold [Ct] < 33) [12-15].

The major goal of this Propagate Network study was to determine the practical utility of an open-access, direct RT-PCR assay [10] via an international, interlaboratory ring trial. The study engaged 10 global sites, including laboratories currently experiencing many of the testing limitations described above, in a series of studies involving a common protocol, common reagents, aliquots of identical pooled clinical samples, and purified nucleic acids, using their existing in-house equipment. Our results suggest that open-access, direct RT-PCR assays are a feasible option for more efficient COVID-19 testing as demanded by the growing pandemic.

## 2. Materials and Methods

### 2.1 Participants

The Propagate Network study was coordinated and partially funded by the international nonprofit HESI as part of its global public health mission and via voluntary contributions of time and effort from the participating partners. Special acknowledgment is given to the University of Washington Virology Laboratory (UWVL) for their efforts to prepare and ship samples for this study and to the University of Vermont Larner School of Medicine for their support in refining the study protocols and recruiting partner laboratories.

Ten laboratories were recruited to participate in the trial for the detection of SARS-CoV-2 RNA from patient nasopharyngeal swabs without RNA extraction using kits provided by UWVL (**Table 1**).

**Table 1.** Institutions and Countries of the Laboratories Participating in the Trial.

| Institution | Country | Role in Propagate Network | Laboratory Role in COVID-19 Testing Regionally |
| --- | --- | --- | --- |
| Department of Pathology, College of Medicine, University of Malawi | Malawi | Project C | Collaborating with government public health authorities to provide testing for local population |
| Bauru School of Dentistry, Department of Biological Sciences, University of São Paulo, Bauru, São Paulo, Brazil | Brazil | Projects A and B | Collaborating with government public health authorities to provide testing for local population and students. Laboratory component of the COVID-19 Diagnostic Network from the University of São Paulo |
| Department of Infectious Diseases, Institute of Tropical Medicine of São Paulo, São Paulo, Brazil | Brazil | Projects A, B, and C | Collaborating with government public health authorities to provide testing for local population and students. Laboratory component of the COVID-19 Diagnostic Network from the University of São Paulo |
| Scientific Platform Pasteur-USP, Universidade de São Paulo, São Paulo, São Paulo, Brazil | Brazil | Projects A, B, and C | Collaborating with government public health authorities to provide testing for local population and students. Laboratory component of the COVID-19 Diagnostic Network from the University of São Paulo |
| Biochemistry Division, National Veterinary Research Institute | Nigeria | Project C | Collaborating with government public health authorities to provide testing for local population |
| Instituto de Ciencias e Innovación en Medicina, Facultad de Medicina Clínica Alemana Universidad del Desarrollo, Santiago, Chile | Chile | Projects A, B, and C | Collaborating with government public health authorities to provide testing for local population |
| Institut de Pharmacologie Moléculaire et Cellulaire, Université Côte d'Azur | France | Projects A and B | Basic science, testing developments |
| LBM Bioesterel | France | Projects A and B | Medical laboratory: processes thousands of samples daily for the south east region of France |
| School of Veterinary Medicine, University of the West Indies | Trinidad and Tobago | Projects A, B, and C | Collaborating with government public health authorities to provide testing for local population, returning residents, and migrants |
| Division of Immunobiology, Department of Medicine, Robert Larner, M.D. College of Medicine, University of Vermont | United States | Projects A, B, and C | Basic science, development of streamlined diagnostic RT-PCR assays for SARS-CoV-2 testing/screening in Vermont, determining how viral RNA load in clinical samples correlates with infectiousness |
| Office of Research and Development, U.S. Environmental Protection Agency | United States | Projects A and B | Not engaged in COVID-19 testing on a regular basis |
| University of Washington Virology Laboratory (UWVL), Virology Division, Department of Laboratory Medicine and Pathology, University of Washington | United States | Projects A and B; lead laboratory responsible for Project A/B sample preparation and dissemination | Processes thousands of samples daily for Pacific Northwest Region and elsewhere in the United States |

Laboratories participated voluntarily and were not offered any compensation for their participation. Due to logistical shipping challenges, which were in large part brought on by the pandemic, samples were unable to be sent to Malawi or Nigeria, underlying the hardships some areas face when testing relies on reagents or materials from other countries.

### 2.2 Ethical statement

The samples generated and disseminated as part of Projects A and B were approved under a waiver of consent by the University of Washington institutional review board (IRB; STUDY00000408). The de-identified samples were determined to be exempt because they were not considered human subjects research due to the quality improvement and public health intent of the work. For Project C, participating laboratories sought the locally appropriate review and permissions for use of de-identified clinical samples as described below.

- **University of West Indies:** Based on the Campus Research Ethics Committee at the University of West Indies, this research met the criteria for Exemption. This decision was made by the chair of the ethics committee, Professor Jerome De Liste.
- **Department of Infectious Diseases, Institute of Tropical Medicine of São Paulo:** Sample use was approved by the local ethics committee (Comissão de Ética para Análise de Projeto de Pesquisa; protocol number CAAE 30419320.7.0000.0068). Informed consent was obtained from all the individuals enrolled in this study.
- **National Veterinary Research Institute, Nigeria:** This work and samples were approved for ethical use within the emergency response to COVID-19 control in Nigeria through rapid laboratory diagnosis. The National Veterinary Research Institute in accordance with the World Organization for Animal Health guidance offered its facility for Public Health Service in Nigeria following activation by the Nigerian Centre for Diseases Control of the Federal Ministry of Health.
- **University of Malawi:** Based on a review by the College of Medicine Research and Ethics Committee (COMREC), no IRB approval acknowledgments were required. This was reviewed by the COMREC administrator, Dr. Lucinda Manda-Taylor, and the compliance officer, Khama Mita.
- **University of Vermont:** This work was approved under a waiver of consent by the University of Vermont IRB (STUDY00000881).
- **Instituto de Ciencias e Innovación en Medicina, Facultad de Medicina Clínica Alemana Universidad del Desarrollo, Santiago, Chile:** The study and publication of its data was approved by a decision made by the Scientific Ethics Committee at the Center of Bioethics at the College of Medicine on 17 March 2021 (as approved by Dr. Marcial Osorio (President of the Scientific Ethics Committee) and Javiera Bellolio A. (Executive Secretary of the Scientific Ethics Committee, College of Medicine, Center of Bioethics) upon review of the document “Admin_Comp_200-53 Requests for Release of Clinical Specimens or Results”, as well as this manuscript. The committee made the statement “It is considered that, given the modality of the study, where the identity is duly protected, and, given the importance from the public health point of view, this project has been approved by the Committee for the publication of the data.” The signed document can be provided upon request.
- Bauru School of Dentistry, Department of Biological Sciences, University of Sao Paulo: Sample use approved by the Institutional Review Board of the Bauru School of Dentistry, University of Sao Paulo (CAAE # 32658720.4.0000.5417)

### 2.3 Study design

The Propagate ring trial consisted of three components. Projects A and B engaged participant laboratories in the analysis of pooled samples disseminated from the lead laboratory (UWVL) for the purpose of evaluating the cross-laboratory performance of the direct method with parallel (identical) samples. Project C characterized the feasibility of the direct method as applied to locally sourced samples collected as part of regional public health testing efforts (**Fig 1**).

**Fig 1.**
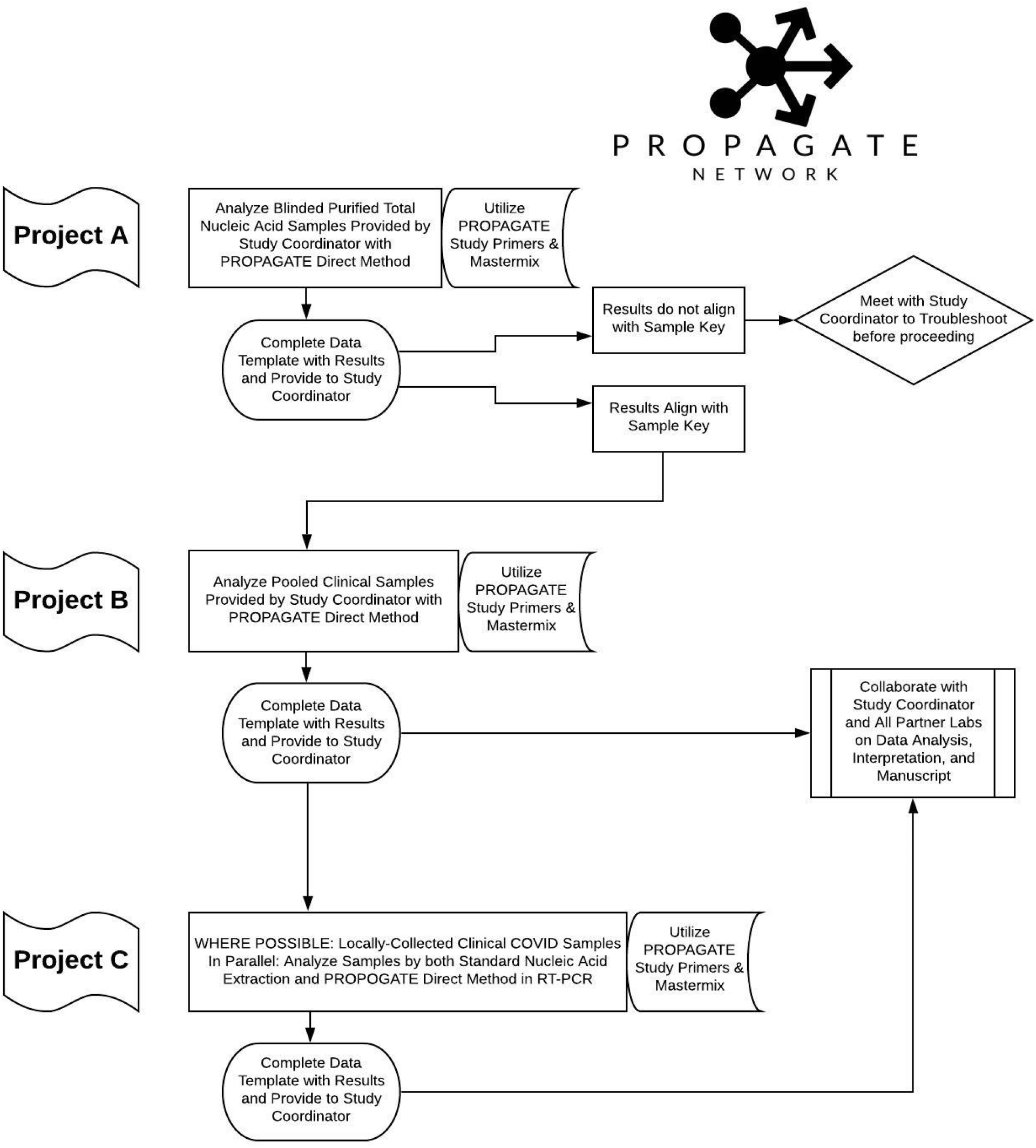
Study Design for the Propagate Network.

All laboratories were invited to participate in Projects A, B, and C. Logistical challenges due to COVID-related shipping restrictions prohibited the involvement of the Malawi and Nigeria laboratories in Projects A and B.

**Project A**. To confirm that all reagents arrived safely and that every laboratory could perform the direct RT-PCR method, each laboratory tested a set of eight nucleic acid samples purified from patients with COVID-19 and supplied by UWVL, which included six blinded samples (three positive and three negative), one sample identified positive, and one sample identified negative to serve as controls, plus a laboratory-supplied no-template water control. Each laboratory reviewed the results of Project A with the study coordinator to confirm that they had correctly identified 100% of the positive and negative blinded samples before proceeding.

Project B. Each laboratory then tested a set of 34 samples supplied by UWVL, including 30 blinded samples (25 positive CT and 5 negative) as well as 2 identified positive and 2 identified negative samples as controls, plus a laboratory-supplied no-template water control. Results for all Project B samples were shared with the study coordinator.

**Project C**. When and where possible, laboratories selected known-positive, locally collected clinical samples and known-negative samples and tested each by both their standard extraction method and the direct RT-PCR method. Results for all Project C samples were shared with the study coordinator.

### 2.4 Heat inactivation

To validate whether a 10-min heating step would inactivate infectious SARS-CoV-2 virus present in clinical samples, the University of Vermont team incubated high-titer stocks of authentic SARS-CoV-2 at 95°C (in a heat block) or room temperature in 1.5-ml Eppendorf tubes for 10 min, spun samples briefly in a microcentrifuge, then measured the infectious units remaining in the heat-treated samples versus the untreated controls by immunofocus assay [16]. The stock virus (strain 2019□nCoV/USA_USA□WA1/2020 [WA1]) was graciously provided by Kenneth Plante and the World Reference Center for Emerging Viruses and Arboviruses at the University of Texas Medical Branch.

### 2.5 Sample preparation

Identical kits were prepared by UWVL and sent on dry ice to each participating laboratory. These kits included reagents for the three possible projects (Fig 1): blinded purified total nucleic acid samples plus positive and negative controls (Project A); blinded unpurified patient samples plus positive and negative controls (Project B); and sufficient enzyme, buffer, primers, and probes to test all samples for Projects A and B, as well as local patient samples (Project C).

To make positive and negative control samples for the kits, nasopharyngeal patient samples in viral transport media were gathered from UWVL’s clinical specimen collection. Three positive samples with a high concentration of SARS-CoV-2 viral RNA (C_t_ ∼15) were pooled together and then diluted 1:32 in a pool of negative samples. Aliquots of positive pooled samples (C_t_ ∼20) and of negative pooled samples were subjected to RNA extraction using a MagNA Pure LC (Roche) to generate purified positive and negative total nucleic acid for Project A.

The unpurified positive pool was diluted further in the negative pool to generate samples at 18 specific expected C_t_ values ranging from 21 to 32 for Project B. These de-identified and pooled samples, both purified total nucleic acid and unpurified patient samples, were each divided into 50-µl aliquots. Two aliquots with a C_t_ value of 21 were included as positive controls in each kit. A random number generator was used to determine the order of blinded samples within the kits. All samples were tested in parallel by direct RT-PCR method and by MagNA Pure LC nucleic acid extraction followed by RT-PCR at UWVL to confirm negative samples and Ct range of positive samples.

### 2.6 Testing method

The direct RT-PCR method is described in Bruce et al. [10]. Briefly, 20 µl of each sample was heat treated for 10 min at 95°C then vortexed and spun down. A Master Mix was made by combining 7 µl of water, 12.5 µl of buffer mix, 1.5 µl of primer/probe mix (IDT), and 1 µl of AgPath-ID enzyme (ThermoFisher) per reaction. Either in 96-well optical PCR plates or optical strip tubes, 22 µl of Master Mix and 3 µl of heat-treated sample was added to each well or tube. All manipulations of clinical samples (transfer for heat inactivation as well as loading of the RT-PCR plate) were performed in a class IIA biosafety cabinet following biosafety level 2 practices. The plates or tubes were then covered with an optical adhesive cover or caps and spun down at 1000 rpm for 1 min. The RT-PCR reaction consisted of 10 min at 48°C for reverse transcription, 10 min at 95°C, and 40 cycles of 95°C for 15 s, followed by 60°C for 45 s with fluorescence measured at the end of each cycle. All samples were tested in duplicate, with water controls on each plate. Reactions to measure the SARS-CoV-2 N-gene (using CDC N2 primers and FAM-labeled probe) and human RNase P gene (using CDC RP primers and FAM-labeled probe) were carried out for each sample in parallel.

### 2.7 Data collection and analysis

For each sample, a mean C_t_ value was computed by averaging individual C_t_ values from all laboratories. A C_t_ value residual (for a given laboratory and sample) was defined as the individual C_t_ value minus the associated mean C_t_ value. For data visualization, individual C_t_ values and residual C_t_ values were plotted against mean C_t_ values. Assay specificity and sensitivity was evaluated using the negative and positive blinded samples.

## 3. Results

### 3.1 Heat inactivation

High-titer stocks of SARS-CoV-2 were treated at 95°C for 10 min. The stock virus had a titer of >10^6^ focus forming units (FFUs) per milliliter. After heat treatment there was more than a 5-log drop, with no detectable foci after 10 min at 95°C (**Fig 2**).

**Fig 2.**
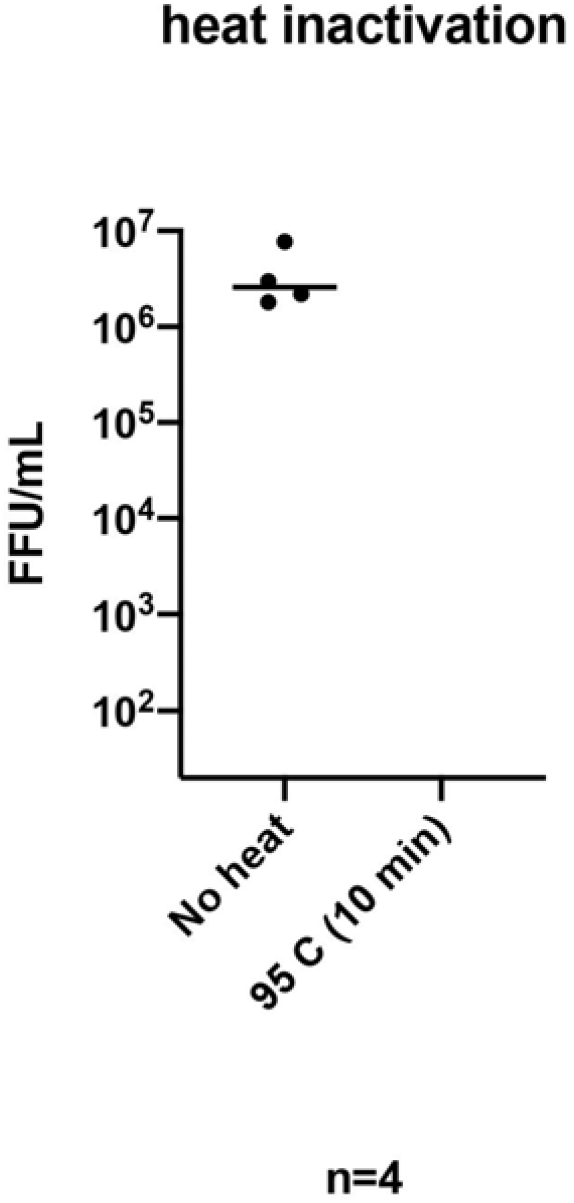
Heat Treatment Inactivates SARS-CoV-2. Stocks of SARS-CoV-2 split into aliquots were treated at 95°C for 10 min or untreated. Viral titer was determined using a focus forming assay with an antibody recognizing the viral N protein. Measurements are in focus forming units (FFU) per milliliter (*n* = 4 replicates). The limit of detection for this assay was 20 FFU/ml.

### 3.2 Project A: laboratory qualification

All participating laboratories correctly identified 100% of the positive (*n* = 3) and negative (*n* = 3) blinded samples sent for purposes of confirming that the samples arrived safely and that the laboratory was able to run the direct RT-PCR method.

### 3.3 Project B: interlaboratory agreement using blinded samples

#### Qualitative agreement between laboratories

The most critical performance measures of a SARS-CoV-2 test are its sensitivity and specificity (simply put, the ability to accurately distinguish the presence versus absence of the viral RNA) and the consistency of its performance across laboratories. As an initial approach, assay specificity and sensitivity was evaluated using 5 known-negative and 25 known-positive samples that were tested in a blinded fashion by the 10 laboratories. For the five negative samples, a total of 50 values were reported by the laboratories, all of which were reported as negative for virus. Thus, the assay demonstrated consistently high (100%) specificity across the laboratories for negative samples (**Table 2**). For the 25 positive samples, the 10 laboratories reported a total of 250 C_t_ values. All of these but one were reported as positive. Thus, all 10 laboratories were able to correctly detect the virus in 24 of 25 samples, and 9 of 10 laboratories were able to correctly detect the virus in all 25 samples using the assay, yielding consistently high [99.6% = (249/250) × 100%] sensitivity across the 10 laboratories for positive samples.

**Table 2.** Specificity and Sensitivity of the Test Based on Results From 30 Blinded Samples (5 Negative and 25 Positive) and the 10 Laboratories.

|  |  | Test |  |
| --- | --- | --- | --- |
|  |  | Negative | Positive |
| Truth | Negative | 50 | 0 |
|  | Positive | 1 | 249 |

#### Quantitative agreement between laboratories

In addition to providing a qualitative determination of the presence versus absence of virus, RT-PCR tests for SARS-CoV-2 can provide additional value by reporting their C_t_ value, which serves as a proxy for the amount of viral RNA present. We therefore investigated the C_t_ values reported for the blinded positive samples tested by the participating laboratories. In general, the agreement between C_t_ values from different laboratories was good, with tighter agreement at lower average C_t_ (higher viral loads) than at higher average C_t_ (lower viral loads; **Fig 3**).

**Fig 3.**
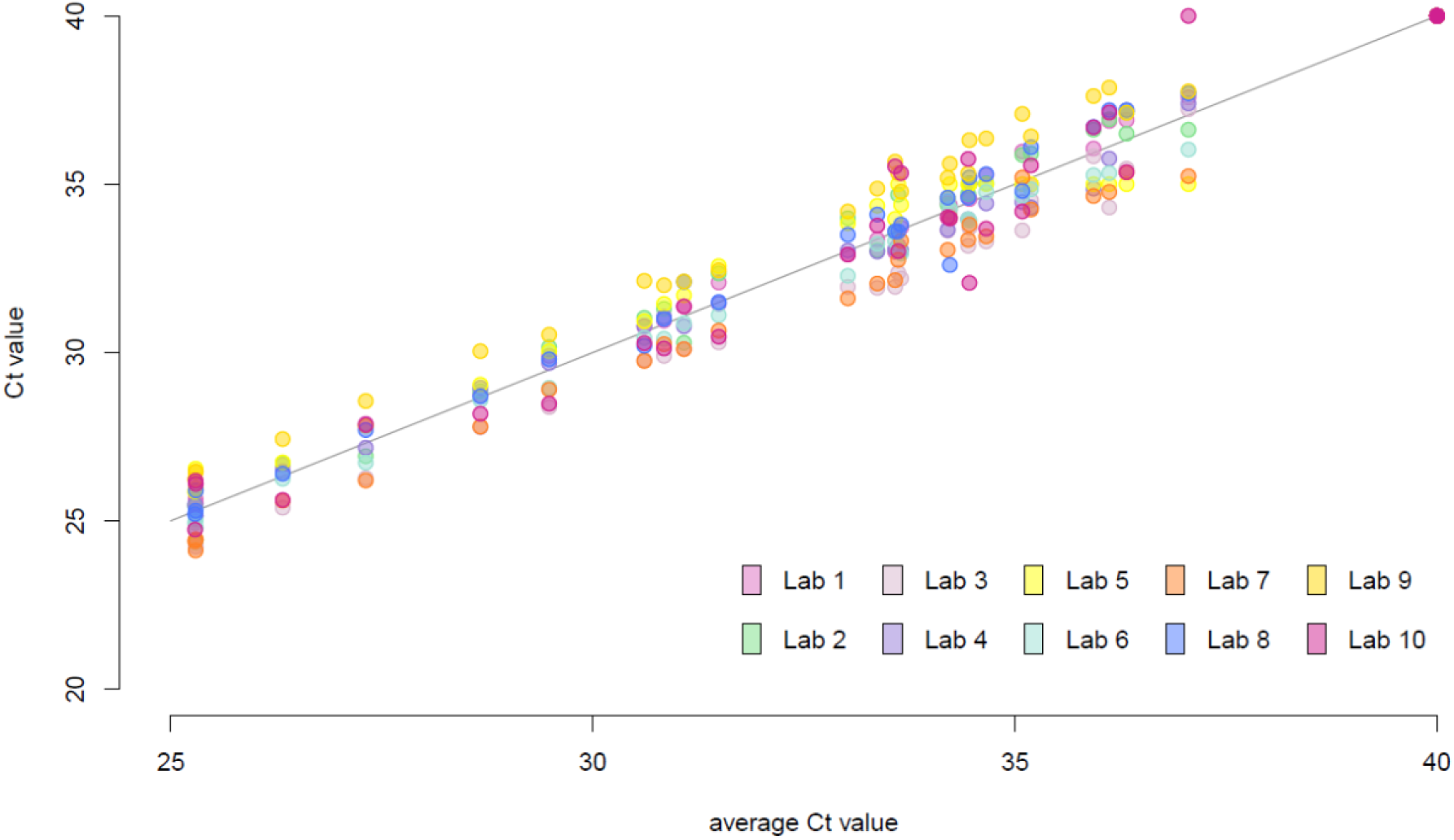
High Interlaboratory Agreement in C_t_ Values for Blinded Clinical Samples in Project B. C_t_ values plotted against the sample-specific average C_t_ values from the 10 laboratories. The solid line indicates the line of equation *y* = *x*. Each color indicates one laboratory; the same color is used to identify each laboratory in all subsequent figures.

We then evaluated the overall quantitative performance of each individual laboratory against the sample-specific average C_t_ value as determined by all 10 laboratories. The C_t_ value residual for a given laboratory and sample was defined as the C_t_ value for the corresponding laboratory and sample minus the sample-specific average C_t_ value; the narrower their distribution within a laboratory, the more consistent the relationship of C_t_ values from the laboratory with the average C_t_ value from all laboratories. Residual C_t_ values had overall similar variability across samples and were minimally affected by the actual viral load (**Fig 4**). The residuals appeared to be centered around zero for most laboratories (**Fig 5**), with the exception of laboratories 3 and 7, for which residuals appeared systematically negative (indicative of C_t_ values consistently lower than average), and laboratory 9, in which residuals tended to be positive (indicative of C_t_ values consistently higher than average). There was no evidence that the assay had a lower sensitivity in this particular laboratory.

**Fig 4.**
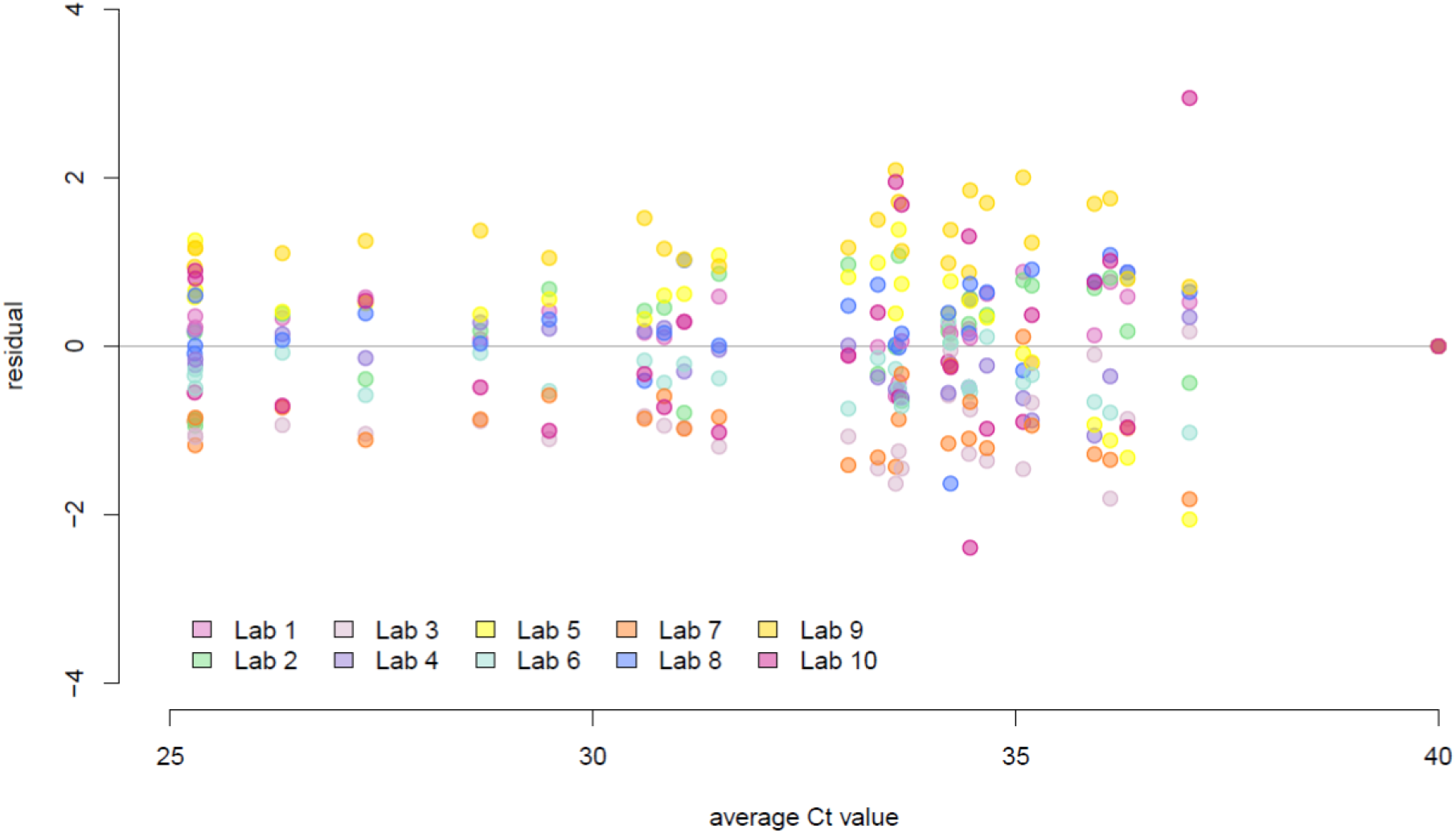
Distribution of C_t_ Residuals Relative to Sample-Specific Average C_t_ Values. Residuals were consistent between laboratories regardless of viral load. Residuals were plotted against sample-specific average C_t_ values. Each colored dot represents a different participating laboratory per legend.

**Fig 5.**
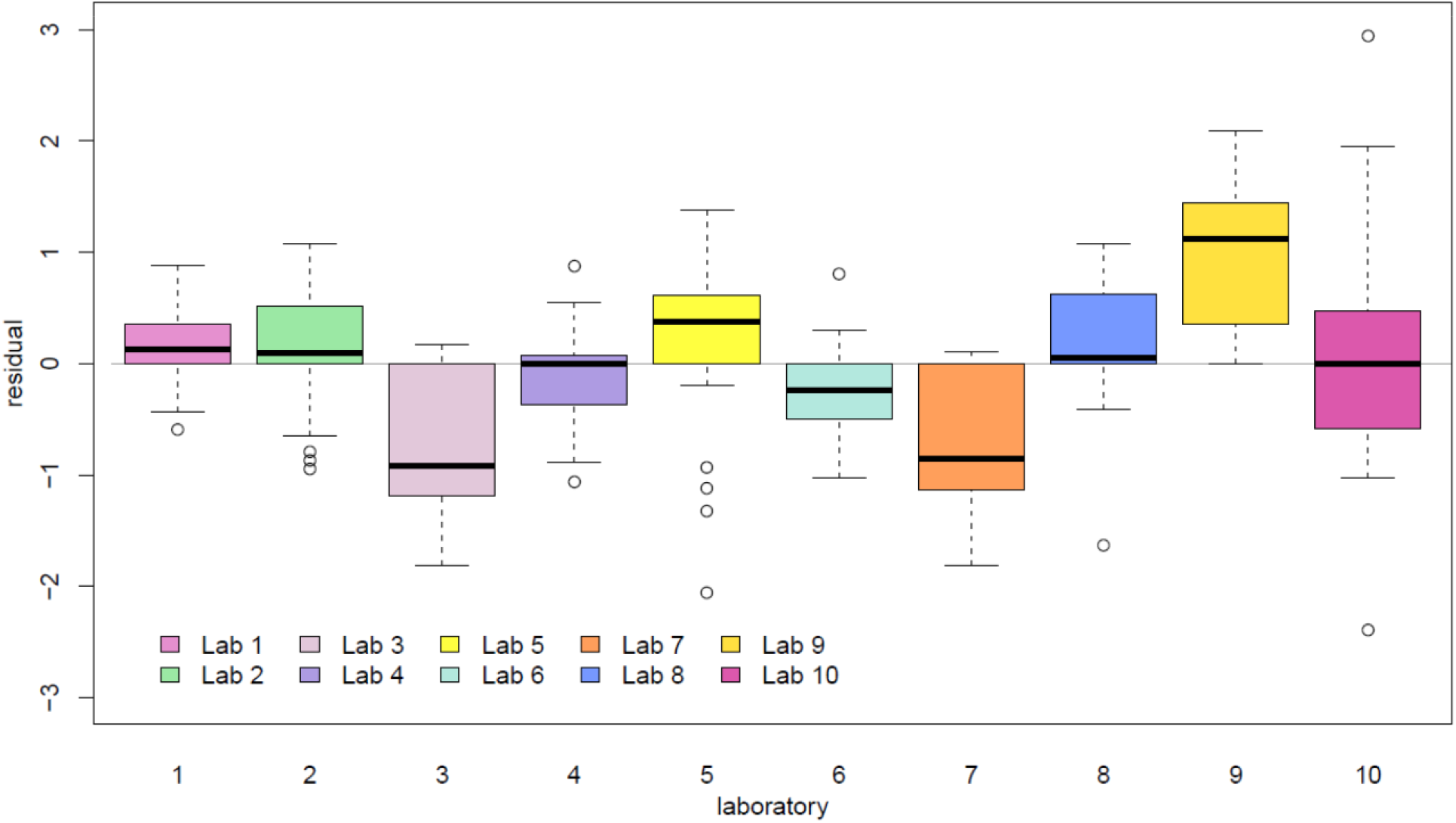
Distribution of Ct Value Residuals Plotted by Laboratory. C_t_ value residuals were calculated as the C_t_ values minus the sample-specific average C_t_ value.

### 3.4 Project C: application of direct RT-PCR on locally collected clinical samples

Seven Propagate partner laboratories conducted side-by-side comparisons of direct RT-PCR and extraction RT-PCR on clinical samples collected from their regions. For four laboratories (2, 7, 8, and 10), these studies demonstrated average losses of sensitivity of between 1.5 and 3.8 cycles in RP C_t_ value (**Fig 6a**; **Table 3**) and between 2.6 and 4.8 cycles in N2 C_t_ value (Fig 6b; Table 3), compared with direct RT-PCR. For RP, this resulted in no failure to detect any sample from any of the four laboratories. For samples in which N2 was detectable by both direct and extraction RT-PCR, the difference in C_t_ values between the two methods did not correlate with the C_t_ value obtained by either method (Fig 6c). However, a few samples (6 of 93) that were detected between C_t_ values of 28 and 39 by extraction RT-PCR were undetectable by direct RT-PCR, while other samples in that range were still detectable by the same laboratories (Fig 6c).

**Table 3.** Difference in Sensitivity of Direct RT-PCR and Extraction RT-PCR Methods on Local Samples From Five Laboratories.

|  | Lab 2 | Lab 7 | Lab 8 | Lab 9 | Lab 10 | All labs |
| --- | --- | --- | --- | --- | --- | --- |
| Samples (*n*) | 20 | 30 | 10 | 4 | 29 | 93 |
| Direct, extracted C<sub>t</sub> (mean ± SD) |  |  |  |  |  |  |
| RP | 2.5 ± 2.5 | 1.5 ± 1.9 | 3.1 ± 0.9 | −0.3 ± 1.4 | 3.8 ± 2.2 | 2.5 ± 2.3 |
| N2 | 4.3 ± 3.2 | 2.6 ± 2.0 | 2.6 ± 1.3 | −1.6 ± 1.4 | 4.8 ± 2.3 | 3.4 ± 3.0 |

**Fig. 6.**
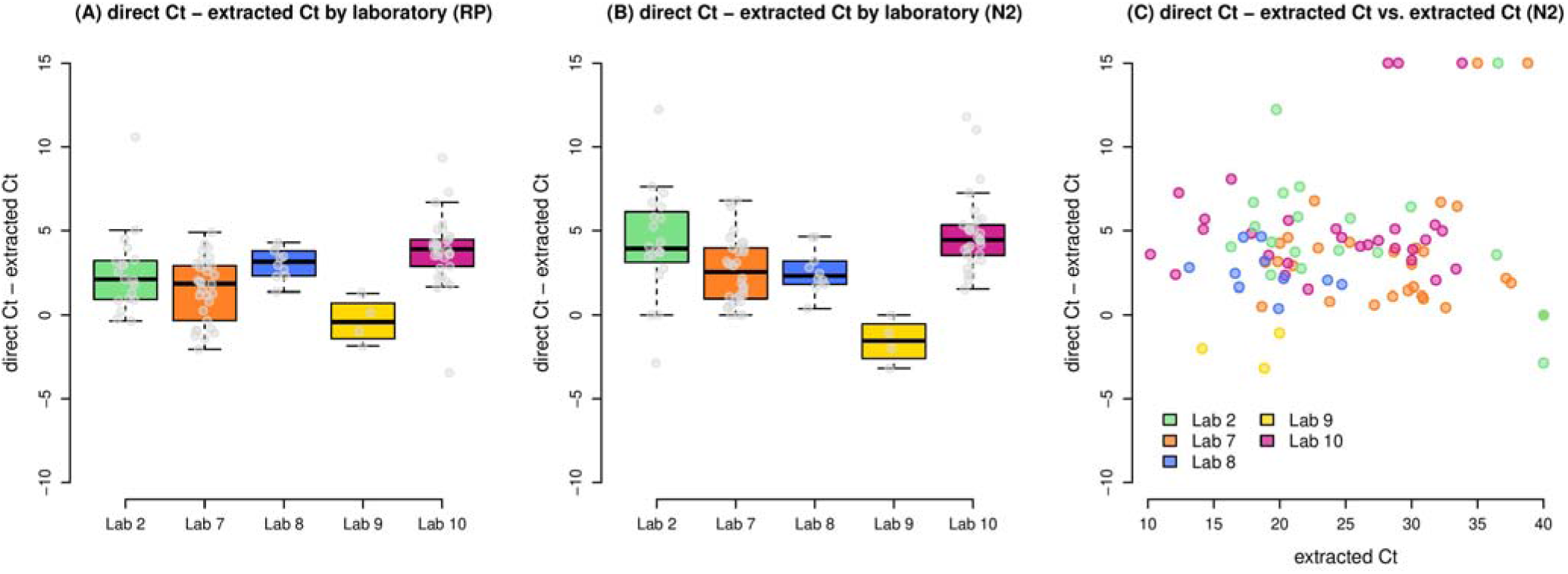
Comparison of Direct Versus Extracted PCR Analyses for Locally Sourced Clinical Samples (Project C). The left (A) and center (B) boxplots illustrate the difference between direct and extracted Ct values in the five laboratories participating in Project C for the RP and N2 genes respectively. The right boxplot (C) illustrates the difference between direct and extracted Ct values plotted against the extracted Ct value, in five laboratories.

For laboratory 9, direct RT-PCR yielded lower Ct values for both RP and N2 than extraction RT-PCR, with an average difference of 0.3 cycles and 1.6 cycles, respectively (Table 3). The reason for this unexpected result is not clear, but it may have been related to the laboratory’s observation that samples became “highly viscous” after the heating step (an observation not reported in any of the other participating laboratories). As with any method, the authors recommend internal validation of the approach prior to clinical implementation.

For two laboratories, direct amplification of both N2 and RP was unsuccessful in all samples, including for samples with low N2 C_t_ values (high viral loads) as measured by extraction RT-PCR. These samples were later determined to have been collected in transport media containing ingredients that were inhibitory to PCR, including charcoal and guanidine (e.g., ManTacc UTM and Jiangsu Rongye Technology LinkGen media were reported as incompatible). Information on the brand/type of viral transport media was not available for all samples used in Project C (this information is often not reported with swabbed samples as provided to analysis laboratories). However, the following media were specifically identified as compatible with this method (Hardy viral transport media, saline, and phosphate-buffered saline).

Overall, the direct approach worked effectively to detect samples deemed positive by standard RT-qPCR when samples were collected in media lacking charcoal or guanine.

## 4. Discussion

This study confirms that the direct RT-qPCR method, initially described by Bruce et al. [10], has the potential to meaningfully contribute to global efforts to detect and contain the COVID-19 pandemic. This study provides evidence that the direct RT-qPCR method is an efficient, reliable, and achievable method for detection of SARS-CoV-2. Although the reproducibility of the method has been reported in single-laboratory studies previously, this study is the first to demonstrate that a globally diverse set of laboratories operating with different equipment, clinical sample collection and handling conditions, resource limitations, and operating practices can successfully implement the method.

As described above, when centrally disseminated pooled samples were evaluated with a common Master Mix and primers/probes (Projects A and B), the Propagate partner laboratory results were >99.5% concordant (all negatives and all but one positive correctly identified and strong agreement on C_t_ values). This result demonstrates the robustness of the methodology. Although all Propagate partner laboratories had prior experience with standard RNA extraction RT-PCR analysis of SARS-CoV-2 test samples, they were able to adopt and implement the direct method on Project A and B samples with only a minimum of instruction (a brief written protocol and a few minutes of discussions via web meeting), showing that the method is easily transferable. Due to halts and delays in air shipments, the partner laboratories in Malawi and Nigeria were unable to receive or analyze the Project A/B sample kits. While this was unfortunate, it is emblematic of the challenges that the African continent (among others) continues to face in receiving needed laboratory supplies and the importance of resource-sensitive methods development efforts such as these.

In Project C of this study, Propagate partner laboratories were encouraged to use their own extraction methods and locally collected samples with RT-qPCR reagents supplied by UWVL to compare results from the direct method versus standard extraction-based PCR. The majority of the participating partner laboratories were successfully able to apply the method and reliably detect RT-PCR-positive samples. The importance of “ground testing” new methods was made evident when some of the laboratories were unable to detect any signal (N2 or RP) following the direct method despite using high-titer positive samples as detected by standard methods. Laboratories experiencing this problem in some cases were working with samples collected in commercial viral transport media that were often found to contain charcoal or in inactivating media such as those containing guanidine. We hypothesize that these are inhibitory to the RT-PCR reaction in the absence of an extraction phase. Similar inhibitory outcomes have been subsequently identified by other laboratories [11, 17]. In other cases, the constituents of media were unknown, so we were not able to hypothesize why the direct method was incompatible. We recommend that laboratories seeking to employ the direct method for SARS-CoV-2 detection should conduct a small pilot run (comparing results from direct and full PCR analyses on the same samples) to ensure that sample media are compatible with this method. This pilot should be replicated if/when sample collection methods or media are changed.

The success of this ring trial is of critical importance given the growing calls for COVID-19 screening as a containment strategy. The growing pandemic requires that we supplement definitive clinical testing with scalable screening strategies that generate efficient, reliable results that can readily inform public health action (e.g., quarantine and isolation) [2]. Non-PCR immunoassay antigen screening kits have decreased sensitivity as compared to standard PCR but are widely utilized depending on the country’s COVID-19 pandemic testing strategy [18]. As anticipated per previous studies, the direct method as applied to SARS-CoV-2 results in some loss in sensitivity compared to standard PCR. One primary explanation for this observation is that RNA extraction typically concentrates RNA present in the clinical sample (by eluting the sample in a smaller volume. In addition, there is a low level of inhibition seen in clinical NP samples loaded directly into an RT-PCR reaction, and the sensitivity of the approach drops when more than 3 ul of patient sample is used [10]. However, this loss is of lower significance to the method’s potential value as a public health screening tool. The direct method succeeds in all of the areas of greatest contemporary need: 1) it reliably detects samples with RNA levels correlating to the presence of live virus (and thus most potential for infectivity), 2) it provides the potential to optimize throughput and reduce costs/logistics for SARS-CoV-2 testing, 3) it is an open-access methodology with no commercial barriers or de novo equipment investment hurdles, and 4) it can be readily adopted by most current public health or clinical laboratories with experience handling infectious samples [14, 15].

We believe that the direct RT-qPCR method for SARS-CoV-2 screening is ripe for adoption in laboratories seeking to reduce turnaround time for processing samples, experiencing challenges in accessing extraction reagents, seeking to decrease costs, and/or looking to reduce the use, handling, and disposal of chemicals in their laboratory. We do not propose this method as a substitute for samples requiring ultrasensitive detection. As with the adoption of any new method, appropriate validation must be conducted by the host laboratory. As standard RNA extraction reagents for PCR can cost $5–$6 USD per extraction and millions of these tests are performed each day around the world, the potential savings are significant. The utilization of this method could lead to greater testing coverage of individuals per dollar invested, or alternatively a larger number of examinations per individual, either of which would allow for the follow up of suspected cases.

The opportunity and feasibility described here is not simply theoretical. At the time of publication, several of the Propagate Network partner laboratories (Brazil, France, United States) are promoting or exploring the broad-scale adoption and implementation of this method for ongoing SARS-CoV-2 public health screening efforts in their regions [19]. Additionally, in October 2020, the Infectious Disease Diagnostic Laboratory at the Children’s Hospital of Philadelphia implemented an extraction-free protocol for routine diagnostic testing of SARS-CoV-2 (*personal communication*). In the 3 months following implementation, >40,000 samples were tested using this workflow. The laboratory observed several critical advantages with this approach, including dramatically reduced extraction reagent costs and a halving of the average laboratory turnaround time, despite increasing test volumes. Further, the independence from specialized extraction reagents for routine testing alleviated pressure on supply chains to meet the increased demand. These same positive impacts on testing efficiency are expected to apply to other laboratories that adopt the method.

While no current testing or screening method is optimal to all situations, the direct method should be considered as a viable, fit-for-purpose resource to address the growing need for population monitoring during a challenging vaccination rollout and amidst the emergence of increasingly virulent strains of SARS-CoV-2.

In addition to the valuable data described above, the global viral testing network established for this study exemplifies the feasibility and importance of establishing transparent, open-access engagements in the public health sciences. Following this study, the Propagate Network will continue to serve as a forum for scientific information exchange and collaboration in the face of future pandemics or health challenges.

## 5. Conclusions

The need for testing for SARS-CoV-2 continues and in many regions is increasing dramatically. This study provides multisite evidence that the direct RT-PCR method can be employed for the detection of SARS-CoV-2 viral RNA with the omission of the RNA extraction step and its associated extraction reagents. This effort represents the first step toward simplifying detection of SARS-CoV-2 viral RNA for the global research community by leveraging evidence-based guidance such as the results present herein. Many options for detecting SARS-CoV-2 have emerged recently, such as antibody testing, saliva testing, and point-of-care testing, which taken together support the urgent need for actionable viral testing. This work lays the foundation for an adoptable method for future viral outbreaks.

## Data Availability

Data can be provided from authors upon request.

## Abbreviations

CDC: Centers for Disease Control and Prevention
COVID-19: coronavirus disease
C_t_: cycle threshold
FFU: focus forming unit
HESI: Health and Environmental Sciences Institute
IRB: institutional review board
N2: specific PCR target within SARS-CoV-2 nucleocapsid (N) gene
RNA: ribonucleic acid
RP: human RNase P gene
RT-PCR: real time–reverse transcription polymerase chain reaction
RT-qPCR: reverse transcription–quantitative polymerase chain reaction
SARS-CoV-2: severe acute respiratory syndrome coronavirus 2
UWVL: University of Washington Virology Laboratory

## Acknowledgments

The authors thank and acknowledge Vianney Leclercq (LBM BIOESTEREL, Mouans-Sartoux, France) for assistance in sample analysis and Bob Bruneau (University of Washington, USA) for coordinating international shipping of samples and reagents. The views expressed in this article are those of the author(s) and do not necessarily represent the views or the policies of the U.S. Environmental Protection Agency.

